# Exposure-Based Video Therapy for Obsessive-Compulsive Disorder and Posttraumatic-Stress Disorder: Clinical Outcomes from a Large Real-World Sample of Adults

**DOI:** 10.1101/2025.11.10.25339930

**Authors:** Nicholas R. Farrell, Clare C. Beatty, Andreas Rhode, Mia Nuñez, Patrick B. McGrath, Larry Trusky, Stephen M. Smith, Jamie D. Feusner

**Author notes:** These authors contributed equally to this work (Nicholas R. Farrell, Clare C. Beatty). **Corresponding Author**: Nicholas R. Farrell, Ph.D., NOCD, Inc., 225 Michigan Ave Ste 1430, Chicago, IL 60601.

## Abstract

**Objective:** Obsessive-compulsive disorder (OCD) and posttraumatic-stress disorder (PTSD) are often comorbid. While exposure-based treatments can be effective for each, given that comorbid OCD and PTSD symptoms often interact dynamically, concurrent treatment may be most beneficial. The aim of the current analysis was to examine the naturalistic effectiveness of concurrently-delivered exposure and response prevention (ERP) and prolonged exposure (PE) therapy among individuals with comorbid OCD and PTSD.

**Methods:** Adult patients (*N*=181) diagnosed with comorbid OCD and PTSD were treated with concurrent ERP and PE as part of a video therapy service specializing in treating comorbid OCD and PTSD. Treatment outcomes for both conditions were assessed at three timepoints: (1) at session 20, (2) at session 40, and (3) the final assessment timepoint completed by each patient.

**Results:** At all three timepoints, there were significant reductions in OCD and PTSD symptoms. By the final timepoint, median percent improvement was 40.9% [IQR: 11.5-70.7%] for PTSD and 50.0% [IQR: 25-67.6%] for OCD. By the final timepoint, 67.4% of patients met criteria for a full PTSD response, 64.1% met criteria for a full OCD response, and 49.2% met criteria for a full response for both conditions. Analyses of secondary treatment outcomes showed significant reductions in depressive and anxiety symptoms, disability, and showed improvements in quality of life.

**Conclusion:** Concurrent ERP and PE, delivered remotely via video therapy in a real-world setting, appears to be a clinically effective treatment approach for individuals with comorbid OCD and PTSD.

## Introduction

With lifetime prevalence rates of approximately 2% and 7% of the general population respectively, obsessive-compulsive disorder (OCD)^1^ and posttraumatic-stress disorder (PTSD)^2^ are common and debilitating. They frequently co-occur with other disorders^1,3^, and with each other: 20–25% of individuals with OCD meet PTSD criteria at some point^4,5^, and comorbid OCD among those with PTSD may be even higher^,4,6^, with one study reporting that 41% of individuals diagnosed with PTSD meet diagnostic criteria for OCD at some point in their lifespan^7^.

In light of the frequent comorbidity of OCD and PTSD, the co-occurrence of these conditions presents notable challenges to symptom alleviation. Their symptoms often interact, worsening distress and complicating treatment (e.g., trauma reminders may trigger safety fears or obsessional doubt about whether the traumatic event occurred)^8^. Most individuals with both disorders (89%) report this dynamic interplay, which is linked to greater overall severity^9^. Compared to either disorder alone, comorbidity is associated with higher symptom burden, poorer quality of life, and more additional mental health conditions^10–12^. Accordingly, treatment studies have documented poorer outcomes among individuals with both OCD and PTSD,^13,14^ and a study found that comorbid PTSD significantly attenuated OCD treatment response^15^. Together, these findings indicate that comorbid OCD and PTSD pose substantial challenges to achieving optimal treatment outcomes.

There have been recent proposals that comorbid OCD and PTSD may be best treated concurrently (i.e., within the same course of therapy)^15, 17^. The two conditions are conceptually similar, both characterized by intrusive thoughts, heightened threat perception and avoidance of distressing stimuli. As noted earlier, most individuals with both disorders report a dynamic symptom interaction, where the same trigger can evoke obsessive-compulsive fear and posttraumatic stress (e.g., public transportation eliciting contamination fears and traumatic memories)^9^. Individuals with both disorders often benefit less from treatments targeting only one condition, partly because symptoms of one can interfere with the other (e.g., in-vivo exposure provoking trauma recollections)^4^. This may explain why comorbid PTSD predicts reduced treatment response among individuals receiving evidence-based psychotherapy for OCD^15^.

To address this, integration of two exposure-based therapies—exposure and response prevention (ERP) and prolonged exposure (PE)— has been proposed for treating comorbid OCD and PTSD^16^. ERP has a strong evidence base, demonstrating it as one of the most effective treatments for OCD, whether psychological, pharmacological, or otherwise^17,18^. Similarly, PE is a gold-standard therapy for PTSD, supported by numerous trials showing significant symptom reduction and quality-of-life improvement^19^. Conceptually, both involve systematic exposure to feared stimuli and elimination of maladaptive coping, such as avoidance, compulsions (for OCD), or safety behaviors. Despite clear theoretical rationale, little is known about their concurrent application. One small study (N=8) provided preliminary support for combined ERP and PE^20^; however, it was limited to an intensive hospital setting.

Importantly, access to ERP and PE has historically been limited by geographical barriers, insufficient numbers of trained providers, and cost—factors contributing to the implementation gap for evidence-based treatments in OCD and PTSD. Recent advances in remote video therapy delivery offer a promising solution, with comparable effectiveness demonstrated for virtual delivery of both ERP for OCD^21^ and PE for PTSD^22^ compared with traditional face-to-face therapy. Teletherapy thus represents a major step toward addressing the longstanding problem of limited access to these treatments^23,24^. However, despite improved availability, no prior studies have examined the combined use of ERP and PE via video therapy for individuals with comorbid OCD and PTSD.

The present observational analysis examined real-world treatment outcomes for adults with comorbid OCD and PTSD who received concurrent ERP and PE. Treatment was delivered via therapist-delivered video therapy on a platform specializing in evidence-based care for both conditions. This retrospective analysis of naturalistic data included patients completing varying numbers and frequencies of sessions.

## Method

### Sample

This retrospective, observational analysis examined clinical data from adults who received combined ERP and PE treatment for comorbid OCD and PTSD through an online specialty therapy platform between July 2022 and July 2025. Patients were first identified through ICD-10 diagnostic codes (F42.x for OCD; F43.x for PTSD) assigned by ERP/PE-trained therapists, yielding 385 potential patients. Inclusion criteria required: (1) clinical diagnoses of both OCD and PTSD; (2) completion of ≥4 sessions each of ERP and PE (n=85 excluded for insufficient PE sessions; n=29 for insufficient ERP sessions); (3) at least two assessments of symptom severity for both disorders (n=21 excluded for insufficient PTSD assessments; n=4 for insufficient OCD assessments); and (4) valid baseline assessments defined as both PCL-5 and DOCS completed within the first two sessions (n=65 excluded). The final analytic sample included 181 patients (M_age_ = 32.0 years, SD = 9.0). Complete demographic and baseline characteristics are presented in Table 1.

**Table 1.** Demographics and psychometrics.

| **Characteristic** | ***n* (%)** | **Mean (SD)** |
| --- | --- | --- |
|  | 181 (100.0) |  |
| **Age** |  | 32.0 (9.0) |
| **Sex** |  |  |
| Female | 146 (80.7) |  |
| Male | 29 (19.9) |  |
| Not reported or prefer not to say | 6 (3.3) |  |
| **Gender<sup>1</sup>** |  |  |
| Cisgender female | 141 (77.9) |  |
| Cisgender male | 23 (12.7) |  |
| Non-binary/gender diverse | 13 (7.2) |  |
| Transgender | 2 (1.1) |  |
| Prefer not to answer | 1 (0.6) |  |
| Missing/unknown | 1 (0.6) |  |
| **Race and Ethnicity** |  |  |
| White | 132 (72.9) |  |
| Hispanic or Latino | 23 (12.7) |  |
| Black/African American | 11 (6.1) |  |
| Asian, Asian American or Pacific Islander | 7 (3.9) |  |
| Other | 4 (2.2) |  |
| Missing/unknown | 2 (1.1) |  |
| Prefer not to answer | 2 (1.1) |  |
| **Pay** |  |  |
| Insurance pay | 167 (92.3) |  |
| Cash pay | 14 (7.7) |  |
| **DOCS** | 181 (100.0) | 35.2 (13.7) |
| **PCL-5** | 181 (100.0) | 43.5 (14.2) |
| **WHODAS (baseline)** | 178 (98.3) | 26.9 (7.8) |
| **QLESQ (baseline)** | 178 (98.3) | 50.4 (14.2) |
| **DASS (baseline)** |  |  |
| Anxiety | 181 (100.0) | 16.6 (8.8) |
| Depression | 181 (100.0) | 17.6 (10.4) |
| Stress | 181 (100.0) | 22.4 (8.7) |
| **Psychiatric Medication<sup>2</sup>** | 110 (60.8) |  |
| SSRI□ | 70 (38.7) |  |
| Anti-anxiety/Sedative-hypnotics □ | 34 (18.8) |  |
| Other Antidepressants □ | 24 (13.3) |  |
| Antipsychotics □ | 21 (11.6) |  |
| Anticonvulsant/Mood Stabilizers □ | 21 (11.6) |  |
| ADHD Medications □ | 16 (8.8) |  |
| SNRI □ | 11 (6.1) |  |
| Tricyclic Antidepressants □ | 8 (4.4) |  |
| PTSD-Specific Medications<sup>i</sup> | 6 (3.3) |  |
| Alpha-2 Adrenergic Agonists<sup>j</sup> | 3 (1.7) |  |
| Sleep Medications<sup>k</sup> | 2 (1.1) |  |
| **Comorbid Diagnoses<sup>2</sup>** |  |  |
| Anxiety<sup>l</sup> | 109 (60.2) |  |
| Mood Disorders<sup>m</sup> | 106 (58.6) |  |
| Somatic Disorders<sup>n</sup> | 15 (8.3) |  |
| Tic Disorders<sup>o</sup> | 12 (6.6) |  |
| Substance Use Disorders<sup>p</sup> | 7 (3.9) |  |
| Eating Disorders<sup>q</sup> | 2 (1.1) |  |
| Adjustment Disorders<sup>r</sup> | 2 (1.1) |  |
| ADHD | 1 (0.6) |  |
| No comorbidities<sup>3</sup> | 33 (18.2) |  |
| Single comorbidity | 63 (34.8) |  |
| Multiple comorbidities | 85 (47.0) |  |

<sup>1</sup>Non-binary/gender diverse includes participants identifying as non-binary (n=6), gender-fluid (n=2), gender-queer (n=2), transgender non-binary (n=2), and other gender identities (n=1).
<sup>2</sup>Some individuals were taking multiple psychiatric medications and/or had multiple comorbid diagnoses; percentages within categories may sum to >100%.
<sup>3</sup>"No comorbidities" refers to participants with PTSD and OCD only, without additional psychiatric conditions. All participants in this study had both PTSD and OCD diagnoses as inclusion criteria.
□ SSRI - Includes fluoxetine, sertraline, escitalopram, citalopram, fluvoxamine
□ Anti-anxiety/Sedative-hypnotics - Includes buspirone, hydroxyzine, clonazepam, propranolol, alprazolam, lorazepam
□ Other Antidepressants - Includes bupropion, trazodone, mirtazapine
□ Antipsychotics - Includes aripiprazole, quetiapine, risperidone, olanzapine
□ Anticonvulsant/Mood Stabilizers - Includes lamotrigine, gabapentin, topiramate
□ ADHD Medications- Includes amphetamine salts, methylphenidate, lisdexamfetamine, dextmethylphenidate, atomoxetine
□ SNRI - Includes venlafaxine, duloxetine
□ Tricyclic Antidepressants - Includes amitriptyline, imipramine, clomipramine
□ PTSD-Specific Medications - Includes prazosin, naltrexone

### Materials

Clinical assessments included the PTSD Checklist for DSM-5 (PCL-5)^25^ for PTSD symptom severity, the Dimensional Obsessive-Compulsive Scale (DOCS)^26^ for OCD symptom severity, and the Diagnostic Interview for Anxiety, Mood, and Obsessive-Compulsive and Related Neuropsychiatric Disorders (DIAMOND)^27^ for diagnostic evaluation of OCD, PTSD, and comorbidities. Secondary measures assessed depression, anxiety, and stress using the Depression, Anxiety, and Stress Scale; (DASS-21^28^), Quality of Life Enjoyment and Satisfaction Questionnaire-Short Form (Q-LES-Q-SF^29^), and disability and functioning with the World Health Organization Disability Assessment Schedule 2.0 (WHODAS 2.0^30^). Detailed descriptions of all measures and administration procedures are provided in the Supplementary Materials.

### Procedures

#### Therapist Training and Treatment Delivery: see Supplementary Materials Data Processing

Baseline assessments were completed between the first and second treatment session following diagnosis. Outcomes were analyzed at sessions 20 and 40 to capture mid- and extended treatment effects, as well as each patient’s final treatment session. When assessments were missed, a last-observation-carried-forward (LOCF) approach was applied, where a patient’s most recent assessment was used until (if) a new assessment was completed. Detailed data processing procedures are described in the Supplementary Materials.

#### Data Analysis

Treatment parameters—including total PTSD and OCD sessions, duration, intensity (sessions per week), and session composition—were calculated for each patient. Primary and secondary outcomes were analyzed using linear mixed models with treatment timepoint (baseline, session 20, session 40, final session) as a fixed factor and patient as a random factor, utilizing restricted maximum likelihood estimation.

Individual-level changes were calculated by comparing baseline with each follow-up assessment. Percent change was computed for each patient and aggregated across the sample. Treatment response was defined as a ≥10-point PCL-5 reduction^31^ and ≥25% reduction (partial) or ≥35% (full) DOCS reduction^32,33^. Combined response required meeting criteria for both measures simultaneously.

Effect sizes were computed using Hedges’ g for paired samples. Missing-data analyses showed no demographic or baseline differences between patients who did and did not reach later treatment timepoints, supporting the Missing-at-Random assumption (see Supplementary Materials). All analyses were conducted in R.

### Ethical Considerations

These analyses did not require research ethics board review, as they do not meet the definition of Human Subjects Research under 45 CFR 46.102(e). This was a secondary analysis of de-identified clinical data obtained retrospectively, with all procedures compliant with NOCD’s privacy policy and applicable data protection laws.

## Results

### Treatment Delivery and Engagement

Patients engaged in therapist-delivered video sessions addressing both OCD and PTSD at a median frequency of 1.1 sessions per week [IQR: 0.9–1.3; ≈4–6 per month]. They completed a median of 34.0 sessions [IQR: 24.0–56.0; M = 42.3, SD = 24.3] over 34.6 weeks [IQR: 21.7–48.4; M = 38.9, SD = 23.9], comprising a median of 24.0 OCD-focused [IQR: 13.0–40.0] and 11.0 PTSD-focused [IQR: 6.0– 16.0] sessions. Engagement varied: 98.9% (n = 179) completed ≥10 sessions, 87.3% (n = 158) ≥20, 60.8% (n = 110) ≥30, and 42.5% (n = 77) ≥40. Among those reaching milestones, median time to session 20 was 15.1 weeks [IQR: 11.9–19.7] and to session 40 was 30.9 weeks [IQR: 25.0–40.7].

On average, treatment included more OCD-focused than PTSD-focused sessions, particularly early in therapy. OCD sessions comprised 87.0%, 79.5%, and 78.5% of total sessions by sessions 10, 20, and 40, respectively, while PTSD sessions represented 13.0%, 20.5%, and 21.5%. Figure 1 illustrates these patterns, showing consistently integrated yet variable treatment delivery across patients.

**Figure 1.**
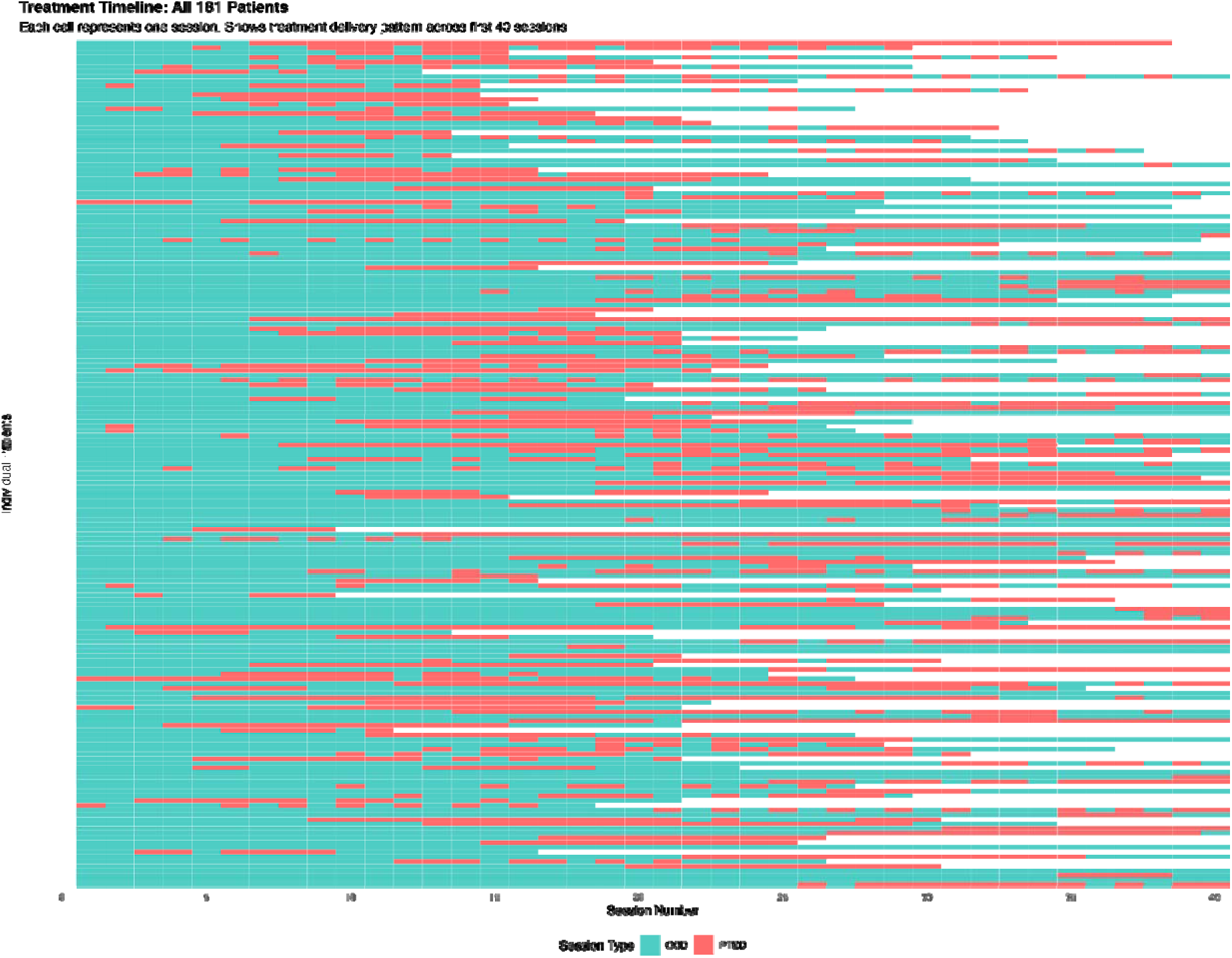
Treatment Session Composition Across First 40 Sessions. Note. Each horizontal row represents one patient’s treatment trajectory (N = 181). Each colored cell represents a single therapy session, with red indicating PTSD-focused sessions and teal indicating OCD-focused sessions. White spaces indicate treatment discontinuation.

### Overall Group Mean Treatment Effects

Across the sample (F□,□.□=39.47, P<.001), PCL-5 scores decreased significantly from 43.5 (SD=14.2) to 35.0 (SD=15.4) at session 20 (−8.5 points, 19.5%; Hedges g=0.56, 95% CI [0.39, 0.73], n=157), and to 33.8 (SD=17.2) at session 40 (−9.7 points, 22.3%; Hedges g=0.64, 95% CI [0.39, 0.89], n=76). Similarly, DOCS scores showed significant improvement (F□,□.□=69.72, P<.001), decreasing from 35.2 (SD=13.7) to 24.5 (SD=14.4) at session 20 (−10.7 points, 30.4%; Hedges g=0.77, 95% CI [0.59, 0.95], n=158), and to 25.4 (SD=15.2) at session 40 (−9.8 points, 27.8%; Hedges g=0.84, 95% CI [0.58, 1.1], n=77) (see Table 2, Figure 2).

**Figure 2.**
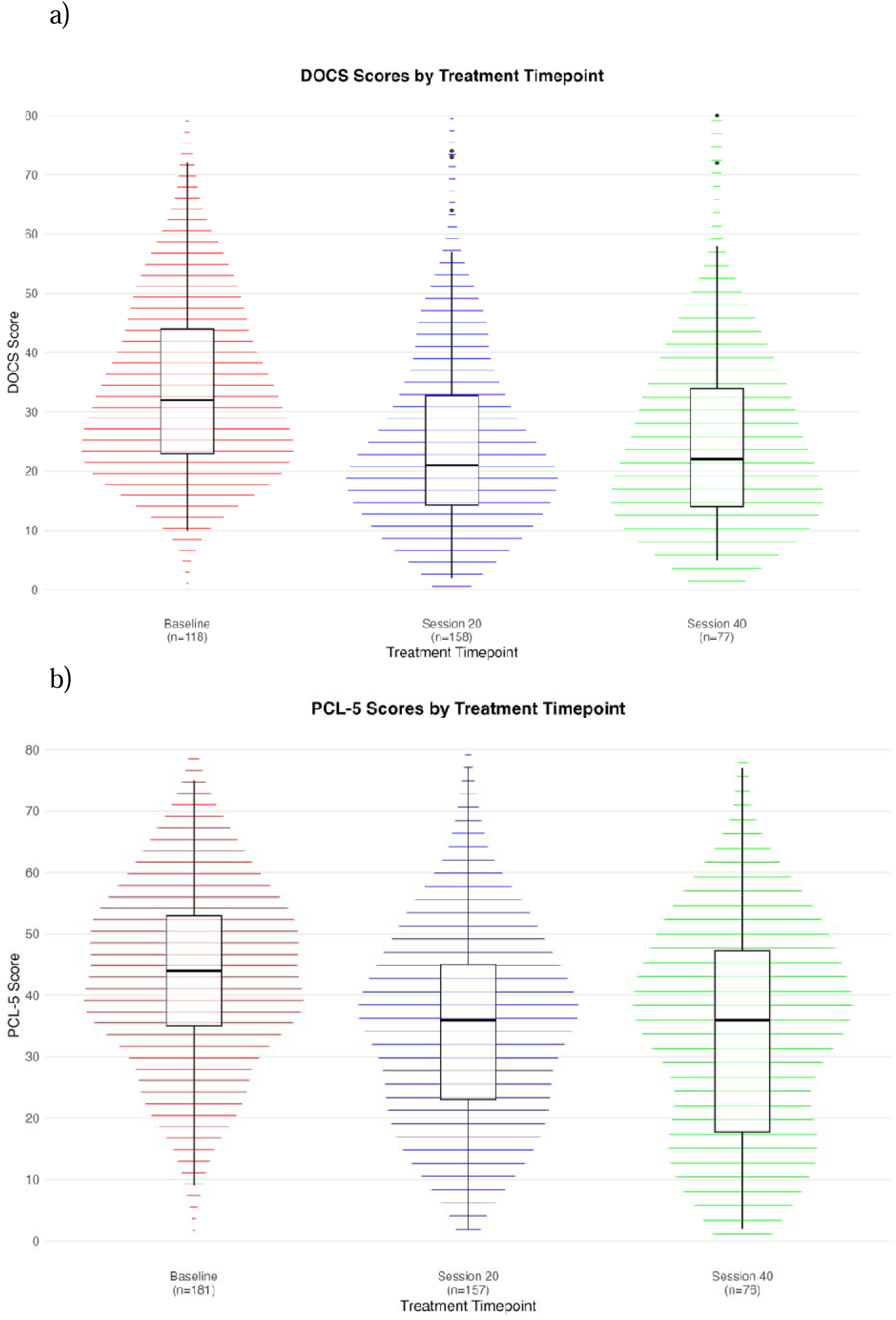
PCL-5 and DOCS Scores by Session. Changes in a) PTSD (top) and b) OCD (bottom) symptoms as assessed by the PCL-5 and DOCS, respectively, with treatment. Median and interquartile ranges are indicated in the box-and-whisker plots. P < .001 for the effect of assessment period. Abbreviations: DOCS = Dimensional Obsessive-Compulsive Scale; PCL-5 = PTSD Checklist for DSM-5.

**Table 2.**
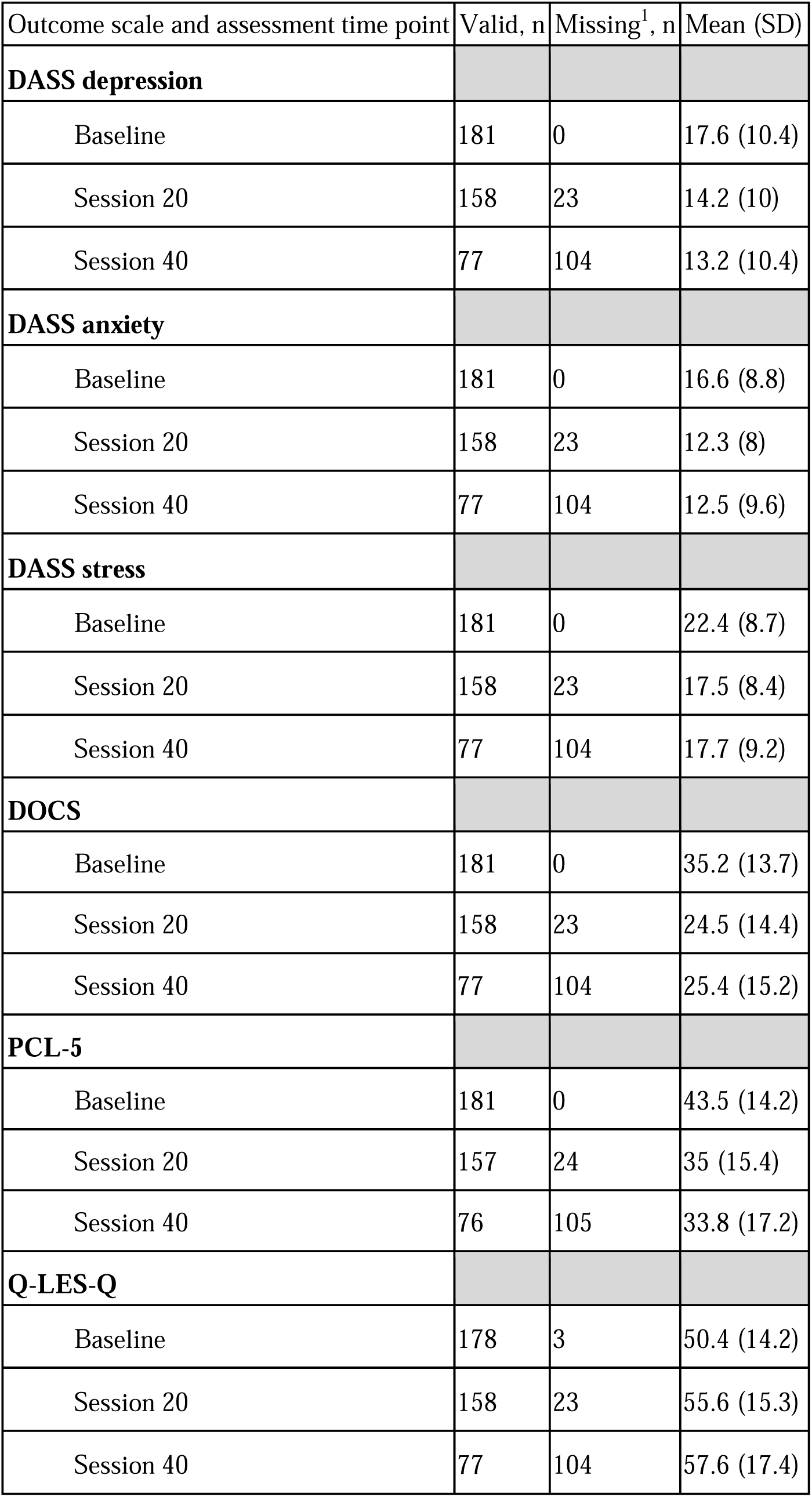
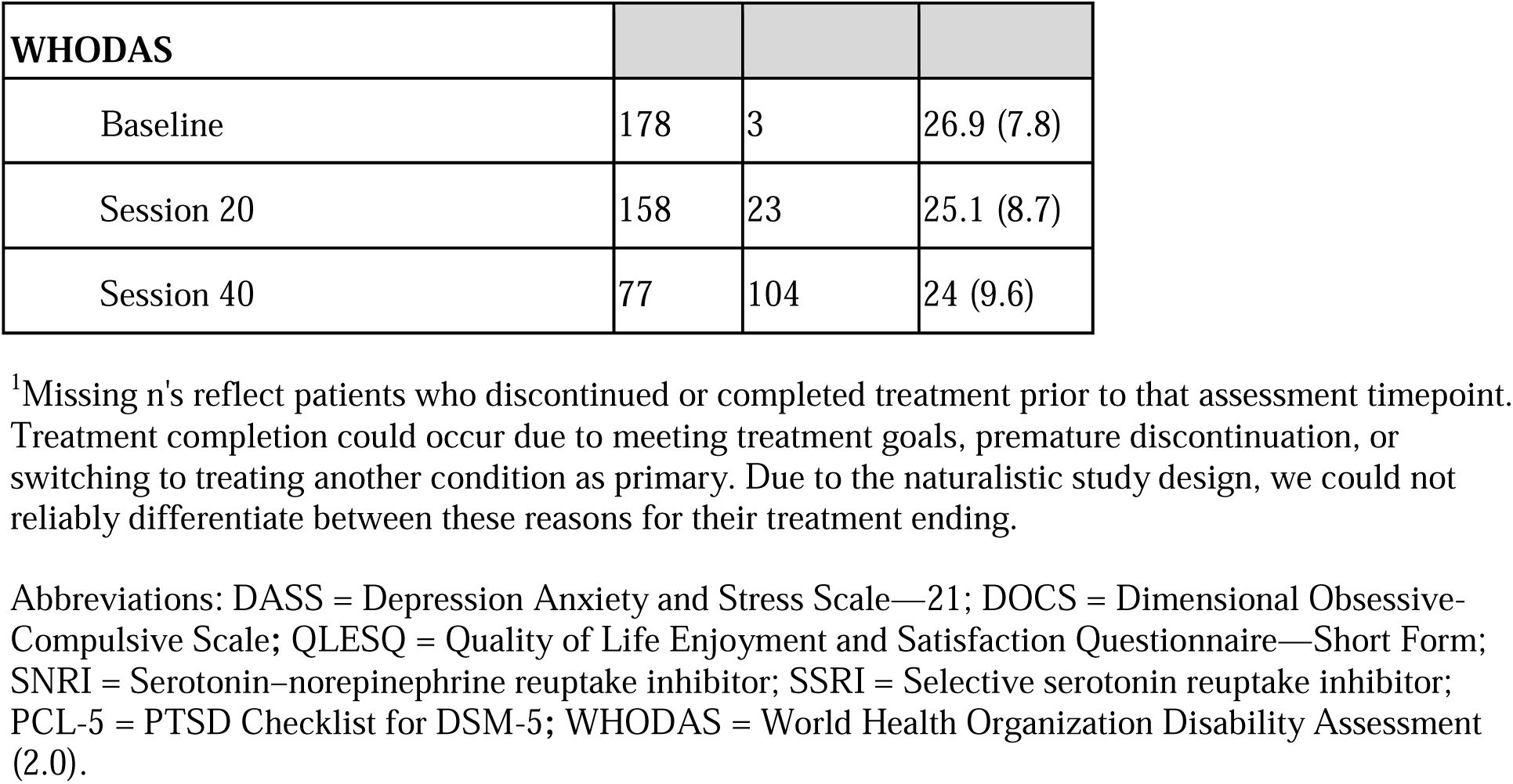
Clinical assessments by treatment time point.

| Outcome scale and assessment time point | Valid, n | Missing<sup>1</sup>, n | Mean (SD) |
| --- | --- | --- | --- |
| **DASS depression** |  |  |  |
| Baseline | 181 | 0 | 17.6 (10.4) |
| Session 20 | 158 | 23 | 14.2 (10) |
| Session 40 | 77 | 104 | 13.2 (10.4) |
| **DASS anxiety** |  |  |  |
| Baseline | 181 | 0 | 16.6 (8.8) |
| Session 20 | 158 | 23 | 12.3 (8) |
| Session 40 | 77 | 104 | 12.5 (9.6) |
| **DASS stress** |  |  |  |
| Baseline | 181 | 0 | 22.4 (8.7) |
| Session 20 | 158 | 23 | 17.5 (8.4) |
| Session 40 | 77 | 104 | 17.7 (9.2) |
| **DOCS** |  |  |  |
| Baseline | 181 | 0 | 35.2 (13.7) |
| Session 20 | 158 | 23 | 24.5 (14.4) |
| Session 40 | 77 | 104 | 25.4 (15.2) |
| **PCL-5** |  |  |  |
| Baseline | 181 | 0 | 43.5 (14.2) |
| Session 20 | 157 | 24 | 35 (15.4) |
| Session 40 | 76 | 105 | 33.8 (17.2) |
| **Q-LES-Q** |  |  |  |
| Baseline | 178 | 3 | 50.4 (14.2) |
| Session 20 | 158 | 23 | 55.6 (15.3) |
| Session 40 | 77 | 104 | 57.6 (17.4) |

| **WHODAS** |  |  |  |
| --- | --- | --- | --- |
| Baseline | 178 | 3 | 26.9 (7.8) |
| Session 20 | 158 | 23 | 25.1 (8.7) |
| Session 40 | 77 | 104 | 24 (9.6) |

<sup>1</sup>Missing n's reflect patients who discontinued or completed treatment prior to that assessment timepoint. Treatment completion could occur due to meeting treatment goals, premature discontinuation, or switching to treating another condition as primary. Due to the naturalistic study design, we could not reliably differentiate between these reasons for their treatment ending.
Abbreviations: DASS = Depression Anxiety and Stress Scale—21; DOCS = Dimensional Obsessive-Compulsive Scale; QLESQ = Quality of Life Enjoyment and Satisfaction Questionnaire—Short Form; SNRI = Serotonin–norepinephrine reuptake inhibitor; SSRI = Selective serotonin reuptake inhibitor; PCL-5 = PTSD Checklist for DSM-5; WHODAS = World Health Organization Disability Assessment (2.0).

### Continued improvement beyond Session 40

From baseline to individually-determined treatment completion (final sessions ranging from 9-121, median=34 [IQR: 24-56]), significant improvement was observed for both PTSD and OCD. PCL-5 scores decreased 17.1 points from baseline (M=43.5, SD=14.2) to final session (M=26.4, SD=17.6) (−17.1 points, 39.3%; F□,□=194.13, P<.001; Hedges’ g=1.03, 95% CI [0.85, 1.21]). DOCS scores also demonstrated significant improvement, decreasing 15.7 points from baseline (M=35.2, SD=13.7) to final session (M=19.5, SD=13.9) (−15.7 points, 44.7%; F□,□=244.55, P<.001; Hedges’ g=1.16, 95% CI [0.97, 1.35]). See Figure 3.

**Figure 3.**
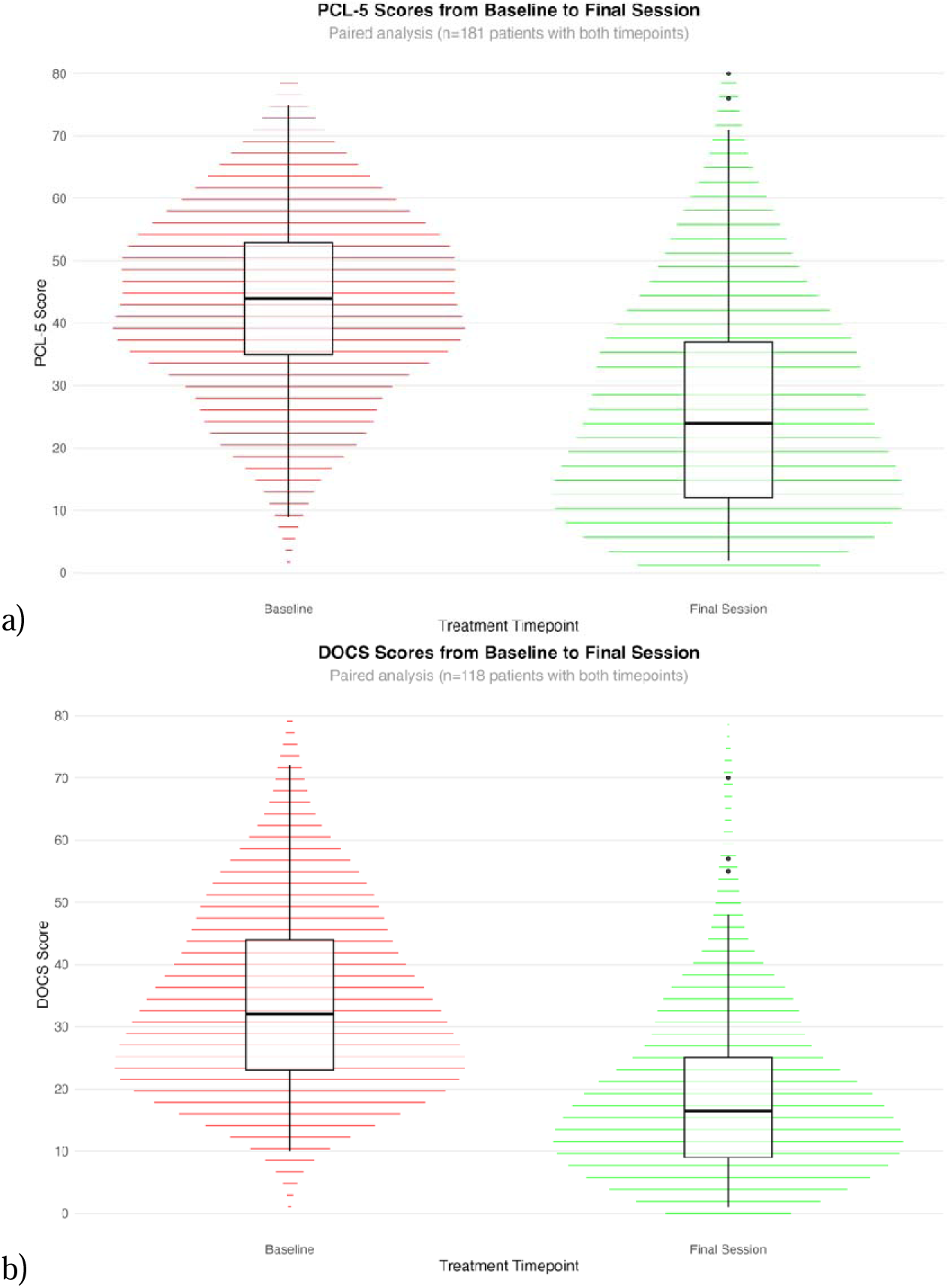
Final Session PCL-5 and DOCS Scores. Changes in a) PTSD (top) and b) OCD (bottom) severity from baseline to final session (range: 9-121 sessions, median: 34 [IQR: 24-56]). Median and interquartile ranges are indicated in the box-and-whisker plots. P < .001 for the effect of assessment period. Abbreviations: DOCS = Dimensional Obsessive-Compulsive Scale; PCL-5 = PTSD Checklist for DSM-5.

### Individual-Level Changes and Response Rates

Median percent improvement was 9.8% [IQR: 0-41.1%] for PTSD and 29.6% [IQR: 3.3-53.8%] for OCD at session 20, and 10.7% [IQR: 0-40.2%] for PTSD and 30% [IQR: 12-49.3%] for OCD at session 40. At session 40, 36.8% (28/76) achieved ≥10-point PCL-5 reduction, 58.4% (45/77) achieved ≥25% DOCS reduction, 42.9% (33/77) achieved ≥35% DOCS reduction, and 17.1% (13/76) achieved combined response (≥35% DOCS and ≥10-point PCL-5 reduction).

At the final session, median improvement reached 40.9% [IQR: 11.5-70.7%] for PTSD and 50.0% [IQR: 25-67.6%] for OCD, with 67.4% (122/181) achieving ≥10-point PCL-5 reduction, 75.7% (137/181) achieving ≥25% DOCS reduction, 64.1% (116/181) achieving ≥35% DOCS reduction, and 49.2% (89/181) achieving combined response (≥35% DOCS and ≥10-point PCL-5 reduction).

### Depression, Anxiety, and Stress, Quality of Life and Disability

Linear mixed models revealed significant improvements across all secondary outcomes from baseline to session 40 (all p<.001). Mean depression, anxiety, and stress scores decreased from 17.6 (SD = 10.4) to 13.2 (SD = 10.4), 16.6 (SD = 8.8) to 12.5 (SD = 9.6), and 22.4 (SD = 8.7) to 17.7 (SD = 9.2), respectively. Functional disability improved from 26.9 (SD = 7.8) to 24.0 (SD = 9.6), and quality of life from 50.4 (SD = 14.2) to 57.6 (SD = 17.4). At the final session, mean percentage improvements were 34.9% for anxiety, 30.1% for depression, 29.9% for stress, 18.8% for quality of life, and 17.5% for disability (all p<.001).

## Discussion

OCD and PTSD frequently co-occur, and individuals with both conditions have historically been at a disadvantage due to interacting symptoms and overall severity. Disorder-specific treatments (e.g., ERP for OCD) may yield suboptimal outcomes when the comorbid condition is not addressed concurrently^15^. Although combining ERP and PE has been theoretically proposed and called for in prior work, few studies have examined its effectiveness for comorbid OCD and PTSD. This analysis aimed to address that gap by evaluating the real-world outcomes of concurrent ERP and PE among individuals diagnosed with both disorders.

Many key outcomes from this analysis indicate that concurrent ERP and PE appears to be an effective approach for individuals with comorbid OCD and PTSD, a relatively common occurrence^4–7^. Patients showed significant reductions in both OCD and PTSD symptom severity. Approximately two-thirds achieved a clinically meaningful (“full response”) reduction in PTSD symptoms, with a similar proportion for OCD, and half met criteria for both. These findings provide some of the first evidence supporting the concurrent use of ERP and PE for this population. Given prior research showing that using only ERP for OCD in such comorbid populations may attenuate treatment response^15^, concurrent application appears to represent a more effective clinical approach.

These findings highlight the need for more clinicians trained in the concurrent use of ERP and PE. Despite their effectiveness, exposure-based treatments are often underutilized or implemented suboptimally, limiting their impact^34^. Many clinicians proficient in one treatment (ERP or PE) lack comparable training in the other^35^, as most specializing trainings focus on disorder-specific rather than comorbid applications. Although transdiagnostic protocols, such as the Unified Protocol for the Transdiagnostic Treatment of Emotional Disorders^36^, has been studied in individuals with OCD and PTSD separately, to our knowledge, it has not been tested specifically for comorbid presentations. Given the dynamic and interactive nature of OCD and PTSD symptoms^9^, clinician training should emphasize managing this complexity within a single, integrated course of ERP and PE rather than delivering them sequentially across providers.

In addition to reductions in OCD and PTSD symptoms, concurrent ERP and PE was associated with improvements in secondary outcomes, including reduced depression and anxiety and increased quality of life and satisfaction. These findings are important, given the high prevalence of depression and anxiety among individuals with OCD and PTSD. Although not directly targeted, these symptoms improved alongside the core disorders, suggesting that treating OCD and PTSD concurrently may alleviate secondary emotional distress.

Another important finding was that many patients continued to experience benefit beyond 40 total sessions. Whereas only 17.1% met criteria for a clinically significant response in both OCD and PTSD at session 40, nearly 50% did so by the final assessment. This suggests that achieving a maximal response to concurrent ERP and PE may require treatment courses longer than those typically used for either disorder alone. Standard ERP averages 12–20 sessions and PE 8–15, so the 20- and 40-session benchmarks were chosen to reflect these ranges. However, the present findings indicate that individuals with comorbid OCD and PTSD may require more extended treatment, as their symptoms often interact dynamically^9^. In practice, clinicians frequently alternated the focus of sessions between OCD and PTSD (see Fig. 1). This may have been necessary when, for example, some OCD-related compulsive behaviors may have additionally served a purpose to manage or prevent intrusive traumatic memories; thus, response prevention in one session may have required prolonged exposure in the next session to address emergent traumatic memories. This treatment pattern may require more sessions than typical single-disorder protocols, as addressing both conditions together could help prevent symptom worsening or attrition. Clinicians applying ERP and PE for comorbid OCD and PTSD should remain flexible and responsive to emerging needs, as many patients may need more than 40 sessions for optimal relief.

The strengths of this analysis include a relatively large sample, examining the effects of concurrent ERP and PE on comorbid OCD and PTSD. As data were drawn from routine clinical care, the findings have strong ecological validity and reflect the expected real-world benefits of concurrent ERP and PE for comorbid OCD and PTSD. Notably, these are the first data to demonstrate the potential effectiveness of concurrent ERP and PE delivered via video therapy rather than in-person sessions. While previous studies have supported ERP for OCD and PE for PTSD individually ^21,22^, provides the first empirical support for their joint use in a video therapy delivery format for individuals with comorbid both conditions.

There are several limitations to this analysis, many related to its observational design. Although all clinicians completed standardized ERP and PE training, no formal fidelity checks (e.g., session video reviews) were conducted, as would occur in clinical trials. However, clinicians participated in biweekly consultation meetings on concurrent ERP and PE delivery, and patient records were routinely audited for accurate documentation. Among the 181 patients, 77 (42.5%) reached session 40, while 98.9%, 87.3%, and 60.8% reached at least 10, 20, and 30 sessions, respectively. For the remaining 104, reasons for not reaching session 40 likely included early treatment completion, shifting focus to other conditions, treatment breaks, discontinuation, or external factors such as insurance or life changes. Although missing data analyses found no baseline differences between those who did and did not reach session 40 (see Supplementary Material), patients who ended treatment earlier may have had different outcomes, potentially leading to overestimation of late-stage effects.

Exposure-based therapies are often believed to be unacceptable and poorly tolerated for many patients,^34^ in spite of research that debunks this concern^37^. However, we did not assess how many patients were offered this treatment but declined, limiting conclusions about acceptability. Additionally, routine post-discharge assessments were not collected, preventing evaluation of long-term maintenance. Finally, outcomes were based on two self-report measures (DOCS and PCL-5), which depend on patients’ ability to accurately distinguish OCD and PTSD symptoms—a challenge noted in prior research^38^. Future work should incorporate clinician- or informant-rated outcomes to complement self-report data.

In sum, concurrent ERP and PE delivered via video teletherapy was associated with clinically meaningful reductions in OCD and PTSD symptoms, along with improvements in quality of life and satisfaction among a large, diverse adult sample. More than half of patients achieved a meaningful response in both conditions, and secondary analyses indicated additional improvements in depression, anxiety, and stress. These results build on previous work with smaller samples,^20^ providing preliminary real-world support for the potential benefits of combining ERP and PE to treat individuals with co-occurring OCD and PTSD.

## Data Availability

The data sets analyzed during this study are proprietary business assets of NOCD Inc., and are not publicly available. The data may be available from the corresponding author on reasonable request with appropriate data use agreements in place.

## Relevant Financial Relationships

NRF, CCB, MN, PBM, LT, SMS, and JDF report personal fees from NOCD Inc.

## Acknowledgments

The authors thank the NOCD therapists, Member Advocates, and the NOCD Operations Team for their help in facilitating treatment and care experiences.

## Supplemental Materials: Method

### Assessments

#### Depression, Anxiety, and Stress Scales (DASS-21)

The DASS-21^1^ is a 21-item self-report measure of symptoms of depression, anxiety, and stress. Patients responded to each DASS-21 item using a scale ranging from 0 (Did not apply at all) to 3 (Applied very much or most of the time) to indicate the extent to which each symptom was experienced during the past week. The DASS-21 factors into three unique subscales measuring depression, anxiety, and stress. In the present analysis, DASS-21 subscale scores were used to assess the effects of the combination of ERP and PE on these secondary treatment outcome variables.

#### Diagnostic Interview for Anxiety, Mood, and Obsessive-Compulsive and Related Neuropsychiatric Disorders (DIAMOND)

The DIAMOND^2^ is a comprehensive semi-structured diagnostic interview that is used to initially screen for and diagnose a variety of mental health conditions in DSM-5. Patients first completed a brief screening questionnaire from the DIAMOND used to identify clinical problem areas warranted further semi-structured assessment. Therapists then administered the semi-structured interview with patients during intake sessions to assess and diagnose OCD, PTSD, and other comorbid conditions. The DIAMOND has previously demonstrated good psychometric properties, including diagnostic reliability, test-retest reliability, and convergent validity.

#### Dimensional Obsessive-Compulsive Scale (DOCS)

The DOCS^3^ is a well-established measure of the nature and severity of OCD symptoms. It is a 20-item self-report questionnaire that uniquely assesses obsessive-compulsive symptoms across four domains that have been consistently found in previous factor analytical research: contamination, responsibility for harm or mistakes, unacceptable thoughts, and incompleteness or symmetry. Patients responded to each DOCS item using a scale ranging from 0 to 4, based on their experiences in the past month. Total scores on the DOCS range from 0 to 80, with higher scores indicating greater symptom severity. The DOCS has shown good psychometric properties, including strong convergent validity with other established measures of OCD symptom severity, and is sensitive to the effects of treatment.

#### PTSD Checklist for DSM-5 (PCL-5)

The PCL-5^4^ is a widely-used measure that assesses the 20 symptoms of PTSD that are listed in DSM-5. It is a 20-item self-report questionnaire that has been used both for diagnostic screening purposes as well as examining the effects of treatment for PTSD. Patients responded to each PCL-5 item using a scale ranging from 0 (Not at all) to 4 (Extremely) to indicate the extent to which each symptom pertained to them during the past month. Total scores on the PCL-5 range from 0 to 80, with higher scores indicating greater PTSD symptom severity. The PCL-5 has shown good psychometric properties, including the ability to detect reliable and clinically significant change in PTSD symptom severity^5^.

#### Quality of Life Enjoyment and Satisfaction Questionnaire-Short Form (Q-LES-Q-SF)

The Q-LES-Q-SR^6^ is a 14-item self-report assessment of quality of life, enjoyment, and overall satisfaction across a variety of life domains. It has demonstrated good psychometric properties in previous research.

#### World Health Organization Disability Assessment Schedule (WHODAS-2.0)

The WHODAS-2.0^7^ is a 12-item self-report questionnaire that assesses difficulties that may be experienced by patients to varying degrees in relation to a health condition. Examples of the difficulties assessed include challenges with taking care of household responsibilities and other activities of daily living, such as hygienic routines and dressing oneself. The WHODAS-2.0 has been used in previous research and demonstrated good psychometric properties.

### Procedures

#### Therapist Training and Treatment Delivery

Therapists who provided treatment completed extensive training programs in ERP for OCD and PE for PTSD. These training programs each lasted approximately 30 hours and both primarily focused on the following areas: (1) the nature and assessment of the condition, (2) conceptualization of the condition with emphasis on maintaining factors, (3) patient-focused psychoeducation, (4) treatment planning via development of exposure hierarchies, and (5) completion of exposure activities within and between therapy sessions. Although the majority of the training was provided in a series of traditional didactic seminars, there were also various experiential learning/training activities, including group discussions of training content and practice (i.e., mock) therapy sessions with clinical experts role-playing as “patients.” Therapists were required to successfully complete these mock therapy sessions with a passing score before they were eligible to begin working with patients. Upon completion of these training programs, therapists were eligible to begin providing ERP and PE to patients. Therapists were required to attend biweekly group case consultation meetings focused on treatment of OCD with ERP and PTSD with PE.

When primarily focused on treating OCD, therapists were trained to recommend and schedule two 60-minute ERP sessions per week until achieving a clinically significant reduction in symptoms, which is consistent with previous work supporting the use of starting with two ERP sessions per week for OCD^8^. For details of the delivery of ERP for OCD, please see the previously-described materials^8^.

When primarily focused on treating PTSD, therapists encouraged their patients to complete one 60-minute PE session per week, consistent with the recommended frequency of PE sessions^9^. Therapists were trained and supervised to encourage continuation of weekly PE sessions until the patient had achieved a clinically meaningful reduction in PTSD symptoms, at which time the therapist and patient either agreed to end PE or reduce the frequency and/or duration of PE sessions to support the patient’s maintenance of their symptom relief. The PE treatment model was based on the work of Foa and colleagues^10^ that has amassed a significant degree of scientific support for its efficacy in treating PTSD among a diversity of populations in various clinical settings^8^.

All sessions were conducted via Zoom (US Health Insurance Portability and Accountability Act–compliant version). Patients were able to join their session from any computer or personal electronic device with access to the Internet. Therapists and patients were required to be on video throughout the duration of all sessions for billing purposes. Throughout the course of concurrent ERP and PE, patients were able to exchange messages with their therapist via the program software used. Patients also had consistent access to therapist-led support groups as well as an online community.

### Data Analysis

#### Initial Sample Refinement

Before episode selection, we applied initial filtering to ensure patients received both treatment modalities. Patients were required to have at least one session with PTSD as the primary diagnosis (F43.x) and at least one session with OCD as the primary diagnosis (F42.x) documented across their entire treatment history.

#### Treatment Episode Detection and Selection Algorithm

Episodes were defined using clinical thresholds: gaps >180 days between consecutive PTSD/OCD sessions indicated the start of a new treatment episode. We selected the first episode containing ≥3 sessions, including ≥1 PTSD session for each patient. For 179 patients (98.9%), their chronologically first episode met this criterion. For 2 patients (1.1%), their first episode had insufficient sessions, so we selected their second episode. Selected episodes were then evaluated against inclusion criteria (≥4 sessions each of ERP and PE, adequate assessments, valid baseline) as described in the main manuscript. PTSD/OCD sessions within selected episodes were renumbered consecutively (1, 2, 3…) to create standardized treatment sequences. Treatment often continued beyond selected episode boundaries (see **Treatment Episode)** but outcomes were analyzed only within the identified meaningful treatment periods.

#### Baseline Assessment

Baseline requirements varied by episode type to ensure valid treatment response measurement. Episode 1 users (first treatment episode) required PCL-5 and DOCS assessments completed between sessions 1-2 of their selected episode. Episode 2+ users (subsequent treatment episodes) required fresh baseline assessments, verified by documented increases in assessment completion counts compared to pre-episode levels. This approach ensured that baseline measurements reflected symptom severity at the start of the analyzed treatment period rather than carry-over from previous treatment attempts.

#### Primary Outcomes

Linear mixed models included all patients (N=181) regardless of whether they reached all post-baseline timepoints. For calculations requiring paired observations (effect sizes, percent reductions, and response rates), analyses were limited to patients with data at both baseline and comparison timepoints.

#### Missing Data

Missing data patterns were systematically evaluated by comparing treatment completion groups on demographic and baseline clinical characteristics. Patients were categorized into three completion groups based on their treatment engagement: Early completion (<20 sessions), Session 20 completers (20-39 sessions), and Session 40 completers (≥40 sessions). One-way ANOVA tests examined continuous variables (age, baseline PCL-5, baseline DOCS) and chi-square tests examined categorical variables (gender, race/ethnicity).

#### Treatment Parameters

Treatment metrics were calculated within selected episodes: total PTSD/OCD sessions (by ICD-10 codes F43.x/F42.x), duration (first to last PTSD/OCD session in weeks), intensity (sessions/week), and session composition (PTSD vs OCD proportions).

#### Effect Size

Hedges’ g was calculated using paired-samples approach with small-sample correction: J=1 - (3/(4*df-1)) where df=n-1. Confidence intervals used SE=√[(1/n) + (g²/(2n))].

#### Moderator Analyses

Potential moderators of treatment response were examined using repeated measures linear mixed models with interaction terms (timepoint × moderator) and random intercepts for patients. Five moderators were tested for each primary outcome: baseline symptom severity (baseline PCL-5 for PTSD outcomes, baseline DOCS for OCD outcomes), treatment dose (total sessions), treatment duration (weeks), and treatment focus (proportion of PTSD vs OCD sessions). Cross-domain moderation was also examined (baseline PCL-5 moderating OCD outcomes, baseline DOCS moderating PTSD outcomes). All moderators were mean-centered continuous variables, and analyses used paired baseline to final session data (N=181 patients). Significant interactions were followed up with simple slopes analyses at low, moderate, and high baseline symptom levels relative to the clinical sample distribution (PCL-5 scores of 33, 45, and 65; DOCS scores of 15, 25, and 35) or treatment focus levels (25%, 50%, 75% PTSD focus).

## Supplemental Materials: Results

### Treatment Episode

Selected episodes demonstrated strong PTSD/OCD treatment focus, with mean episode length of 43.1 sessions (SD=24.3) comprising 13.2 PTSD sessions (SD=9.8) and 29.2 OCD sessions (SD=18.1). PTSD sessions represented 30.5% of episode sessions, while OCD sessions comprised 67.7%. Overall, 98.2% of sessions within analyzed episodes targeted PTSD or OCD symptoms.

Post-episode treatment patterns indicated that selected episodes represented substantial portions of patients’ total PTSD/OCD care. Among all patients, 35.4% continued PTSD/OCD treatment beyond their selected episode after a median gap of 5.9 weeks [IQR: 4.9-7.3 weeks], while selected episodes comprised a mean of 92.2% (median 100% [IQR: 90.9-100%]) of patients’ total PTSD/OCD treatment.

For the 2 patients requiring Episode 2+ selection, pre-episode treatment averaged 1.5 PTSD/OCD sessions (range 1-2), confirming minimal prior exposure to target condition treatment before the analyzed episodes.

### Missing Data

No significant differences were found across completion groups for age, F(2,178)=0.60, p=0.552, baseline PCL-5, F(2,178)=0.12, p=0.890, baseline DOCS, F(2,178)=0.20, p=0.816, or gender distribution (χ²=0.08, p=0.961). These findings support analysis under the Missing At Random (MAR) assumption.

### Moderators

Baseline symptom severity moderated treatment effects for both primary outcomes. For PTSD symptoms, the timepoint × baseline PCL-5 interaction was significant, F(1, 358)=25.25, p<0.001.Simple slopes analyses revealed that patients with higher baseline severity showed greater absolute symptom reduction: baseline PCL-5 of 33 showed 12.8-point improvement (95% CI: 10.0-15.6), baseline PCL-5 of 45 showed 17.7-point improvement (95% CI: 15.4-20.0), and baseline PCL-5 of 65 showed 25.9-point improvement (95% CI: 21.8-30.0). Percentage improvements were similar across severity levels (approximately 39%).

For OCD symptoms, the timepoint × baseline DOCS interaction was also significant, F(1, 358)=54.05, p<0.001. Simple slopes analyses showed absolute improvements of 6.1 points (95% CI: 3.0-9.2) for baseline DOCS of 15, 10.9 points (95% CI: 8.7-13.0) for baseline DOCS of 25, and 15.6 points (95% CI: 13.9-17.4) for baseline DOCS of 35. Percentage improvements remained consistent across severity levels (42-45%).

Treatment focus moderated PTSD outcomes, with a significant timepoint × PTSD proportion interaction, F(1, 179)=5.54, p=0.02. Simple slopes analyses indicated greater PCL-5 improvements with higher PTSD session focus: 15.6-point reduction (95% CI: 12.9-18.3) for low PTSD focus (25%), 19.5-point reduction (95% CI: 16.4-22.7) for balanced focus (50%), and 23.5-point reduction (95% CI: 17.7-29.4) for high PTSD focus (75%).

Treatment dose and duration interactions were not significant for either outcome (all F<1.78, all p>0.18). Cross-domain interactions were also non-significant (all F<0.50, all p>0.75).

